# Transcriptomic profiling of disease severity in patients with COVID-19 reveals role of blood clotting and vasculature related genes

**DOI:** 10.1101/2020.06.18.20132571

**Authors:** Kiran Iqbal Masood, Syed Faisal Mahmood, Saba Shahid, Nosheen Nasir, Najia Ghanchi, Asghar Nasir, Bushra Jamil, Iffat Khanum, Safina Razzak, Akbar Kanji, Zahra Hasan

**Affiliations:** Department of Pathology and Laboratory Medicine, Department of Medicine, The Aga Khan University, Karachi, Pakistan

## Abstract

COVID-19 caused by SARS-CoV-2 manifests as a range of symptoms. Understanding the molecular mechanisms responsible for immuno-pathogenesis of disease is important for treatment and management of COVID-19. We examined host transcriptomes in moderate and severe COVID-19 cases with a view to identifying pathways that affect its progression. RNA extracted from whole blood of COVID-19 cases was analysed by microarray analysis. Moderate and severe cases were compared with healthy controls and differentially regulated genes (DEGs) categorized into cellular pathways.

DEGs in COVID-19 cases were mostly related to host immune activation and cytokine signaling, pathogen uptake, host defenses, blood and vasculature genes, and SARS-CoV-2- and other virus-affected pathways. The DEGs in these pathways were increased in severe compared with moderate cases. In a severe COVID-19 patient with an unfavourable outcome we observed dysregulation of genes in platelet homeostasis and cardiac conduction and fibrin clotting with disease progression.

COVID-19 morbidity is associated with cytokine activation, cardiovascular risk and thrombosis. We identified DEGs related to dysregulation of blood clotting and homeostasis, platelet activation pathways and to be associated with disease progression. These can be biomarkers of disease progression and also potential targets for treatment interventions in COVID-19.

## Background

COVID-19 is caused by SARS-CoV-2 which belongs to a family of beta-corona viruses. It was first discovered in Wuhan, a city in the Hubei Province of China in 2019. It belongs to the same subgenus but different clade as the severe acute respiratory syndrome (SARS) virus. COVID-19 is spreading globally but it is not clear as to what are the underlying host immune parameters that determine outcome of the disease. Factors such as, host immune compromised conditions, underlying cardiac disease, kidney disease and advanced age have all be associated with poor COVID 19 outcomes (1, 2).

SARS-CoV-2 attaches via its spike protein to host cells by binding to angiotensin-converting enzyme 2 (ACE2) receptor which is abundantly expressed in the cells of the lower respiratory tract (3, 4). The virus enters via the endocytic pathway, replicates until lyses and spreads to infect neighbouring cells (4). Acute inflammation results from SARS-CoV-2 infection of pneumocytes, which causes a cytopathic effect as it spreads (5). The ‘cytokine storm’ induced via pathways such as the inflammasome (NLRP3) and modulation of host protective innate and adaptive immune cells has a detrimental effect on COVID-19 morbidity and mortality (6). T helper and T regulatory cells which regulate adaptive immunity, are shown to be dysregulated by SARS-CoV-2 infection (7). SARS-CoV-2 also triggers IL-4 and IL-10 which are Th2 cytokines are possible regulators of a homeostatic balance (8). COVID-19 treatment and management involves reducing cellular inflammation through the use of corticosteroids and inflammatory agonists such as to IL-6 (8). As we learn more about disease pathology we can identify appropriate cellular treatments for patient treatment.

Here we investigated the transcriptome profiles of COVID-19 patients with moderate and severe disease compared with healthy endemic controls. We identified differentially regulated genes (DEGs) in a COVID-19 patient with severe disease who expired, to involve blood clotting and homeostasis, platelet and immune activation pathways as those most affected by SARS-CoV-2 infection.

## Methods

This study received approval from the Ethics Review Committee of the Aga Khan University. All COVID-19 patients had a respiratory sample positive for SARS-COV-2 by real-time polymerase chain reaction (RT-PCR) and were admitted at The Aga Khan University Hospital. Healthy endemic controls (EC) were those tested as part of an ongoing study looking at host transcriptome responses in the population. All study subjects were males and females aged over 18 years. Written and /or verbal consent was taken from all study subjects.

There were four COVID-19 cases with a mean age of 57 years. Clinical laboratory parameters depicted (Table 1) were those at the time of recruitment in the study. Samples were taken from patients within 24 – 48 h of admission and prior to intervention with either the IL-6 antagonist Tocilizumab or, steroid treatment. **P1**, had moderate COVID-19; a female (21-35 y) with a history of fever and sore throat prior to presentation and with shortness of breath. She was hypoxic on admission with oxygen saturation of 93%, required oxygen supplementation and showed bilateral chest infiltrates on chest radiograph. She was administered Tocilizumab, subsequently improved and was discharged. **P6**, had moderate COVID-19; a male (51-65 y) who presented with a history of fever and myalgias. He was hypoxic on admission, required oxygen supplementation and showed bilateral chest infiltrates on chest radiograph. He recovered after treatment with Hydroxychloroquine and steroids. **P3**, had severe COVID-19; a (> 65 y) male who was critically ill on admission with Acute Respiratory Distress Syndrome (ARDS), septic shock, acute kidney injury and early disseminated intravascular coagulation (DIC) and required mechanical ventilation. He was treated with Hydroxychloroquine, Tocilizumab and steroids however, he expired. **P5**, had severe COVID-19; a (51-65 y) male who was admitted with ARDS, cytokine release syndrome, acute kidney injury and non-ST elevation myocardial infarction, requiring mechanical ventilation. He responded to Tocilizumab, was extubated, gradually recovered and discharged.

**Table 1.** Clinical characteristics of COVID-19 patients.

| **Specimens** | **Age**<br>**(y range)** | **Gender**<br>**(M/F)** | **Severity** | **D dimer**<br>**(mg/L FEU)** | **CRP**<br>**(mg/L)** | **LDH**<br>**(IU/L)** | **Ferritin**<br>**(ng/mL)** | **NLR** | **POCT-GLUF**<br>**(md/dl)** |
| --- | --- | --- | --- | --- | --- | --- | --- | --- | --- |
| P1 | 21-35 | F | Moderate | 8.9 | 142.59 | 528 | 486.3 | 1.83 | 82 |
| P6 | 51-65 | M | Moderate | 1.1 | 73.83 | 436 | 180.4 | 4.08 | 109 |
| P3-d3 | > 65 | M | Severe | >30 | 209.95 | 821 |  | 19.41 | 147 |
| P3-d6 |  | M | Severe | 18.8 | 99.89 | 615 | 642* | 12.98 | 124 |
| P5 | 50-65 | M | Severe | 3.2 | 201.03 | 354 | 639.54 | 10.77 | 126 |

### Endemic controls (EC)

were healthy individuals with no symptoms of respiratory illness, fever or co-morbid conditions. There 2 females and 2 males with a mean age of 37 y. None of the individuals had co-morbid conditions.

### Laboratory tests

Nasopharyngeal swab samples from patients were tested using the COBAS® SARS-CoV-2 assay (COBAS® 6800 Roche platform). Routine blood hematology and biochemistry tests conducted included the complete blood count, D-dimer, C-reactive protein (CRP), ferritin, lactate dehydrogenase (LDH) and fasting glucose levels.

### RNA microarray analysis

RNA was extracted from whole blood collected in plasma/EDTA tube using the Qiagen RNA Blood Mini Kit (Qiagen, GmbH, Germany). One hundred nanogram of RNA was used for preparation of cRNA for use in the Clariom S Array Type gene expression, Affymetrix. The arrays were scanned using an Affymetrix autoloader system. CEL files were analysed using the TCAS Transcriptome Analysis Software Suite (version 2) using the Summarization Method: Gene Level - SST-RMA Pos vs Neg AUC Threshold: 0.7 against Genome Version: hg38 (Homo sapiens).

### Cellular Pathway analysis

DEGs significantly up- or down-regulated (p<0.05) with Gene fold change < −2 or > 2 were identified by TCAS software and categorised using the WikiPathways. Significantly modified pathways were sub-grouped as; Blood and vasculature; Immune activation and cytokine signaling; Pathogen uptake and host defense; Glucose metabolism; Vesicular transport; Gene regulation; SARS-CoV-2- and other Virus-induced response related genes.

## RESULTS

### Clinical description of study subjects

We studied four COVID-19 cases; one female (P1) and one male (P6) had moderate disease and two males (P3 and P5) had severe to critical disease. All of them displayed increased levels of C-reactive protein (CRP), D-Dimers, lactate dehydrogenase (LDH), Ferritin and raised neutrophil/lymphocyte ratios (Table 1).

### Differential gene expression between controls and COVID-19 cases

We compared the transcriptomic signatures of EC were compared with COVID-19 positive cases (Fig. 1A). Within each comparative set, in EC, 1419 genes were upregulated and 1996 were downregulated as compared with moderate COVID-19 cases (Fig. 1B). As compared with severe COVID-19 cases, 1698 genes were upregulated and 2126 were downregulated in EC. Overall, there were 4958 differentially regulated genes (DEGs) between EC and COVID-19; with 3415 DEGs between EC and moderate cases and 3824 DEGs between EC and severe cases (Fig. 1C). Of these, 2688 DEGs were common between the moderate and severe COVID-19, 702 DEGs were unique between EC and moderate whilst, 1075 DEGs were unique between EC and severe disease.

**Fig 1.**
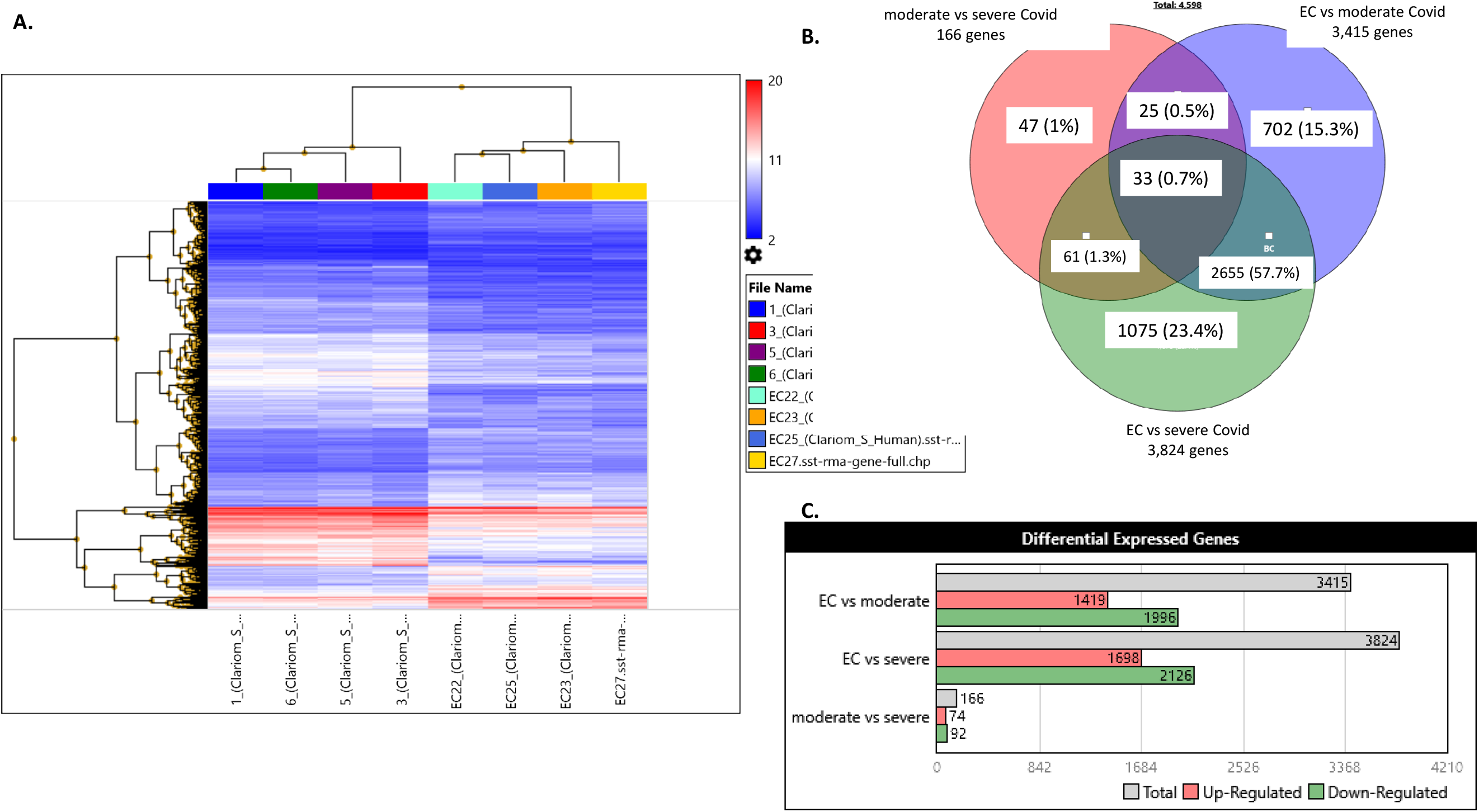
Differential gene regulation between controls and COVID-19 patients with moderate and severe disease. **A**, Heat map showing hierarchical clustering of differentially regulated genes (DEGs) between endemic controls (EC) and Covid-19 patients with moderate or severe disease. B. Venn diagram depicts the overlaps between DEGs observed between the EC and moderate Covid-19, EC and severe Covid-19. **C**.

### Pathways dysregulated in moderate COVID-19 disease

DEGs were identified in host immune activation and cytokine signaling, pathogen uptake and host defense, blood and vasculature, glucose-related metabolism, cell death and repair, vesicular transport and SARS-CoV-2 and other virus-induced pathways. In moderate COVID-19, the greatest number of DEGs were in immune activation and cytokine signaling pathways (70%), SARS-CoV-2 induced pathways and other viral responses (12.2%), glucogenesis related pathways (7.7%), blood and vasculature (6.3%), cell death and repair related genes (5.7%), and those related to pathogen uptake and host defenses (3.3%), Fig. 2A, Supplementary Table 1.

**Fig 2.**
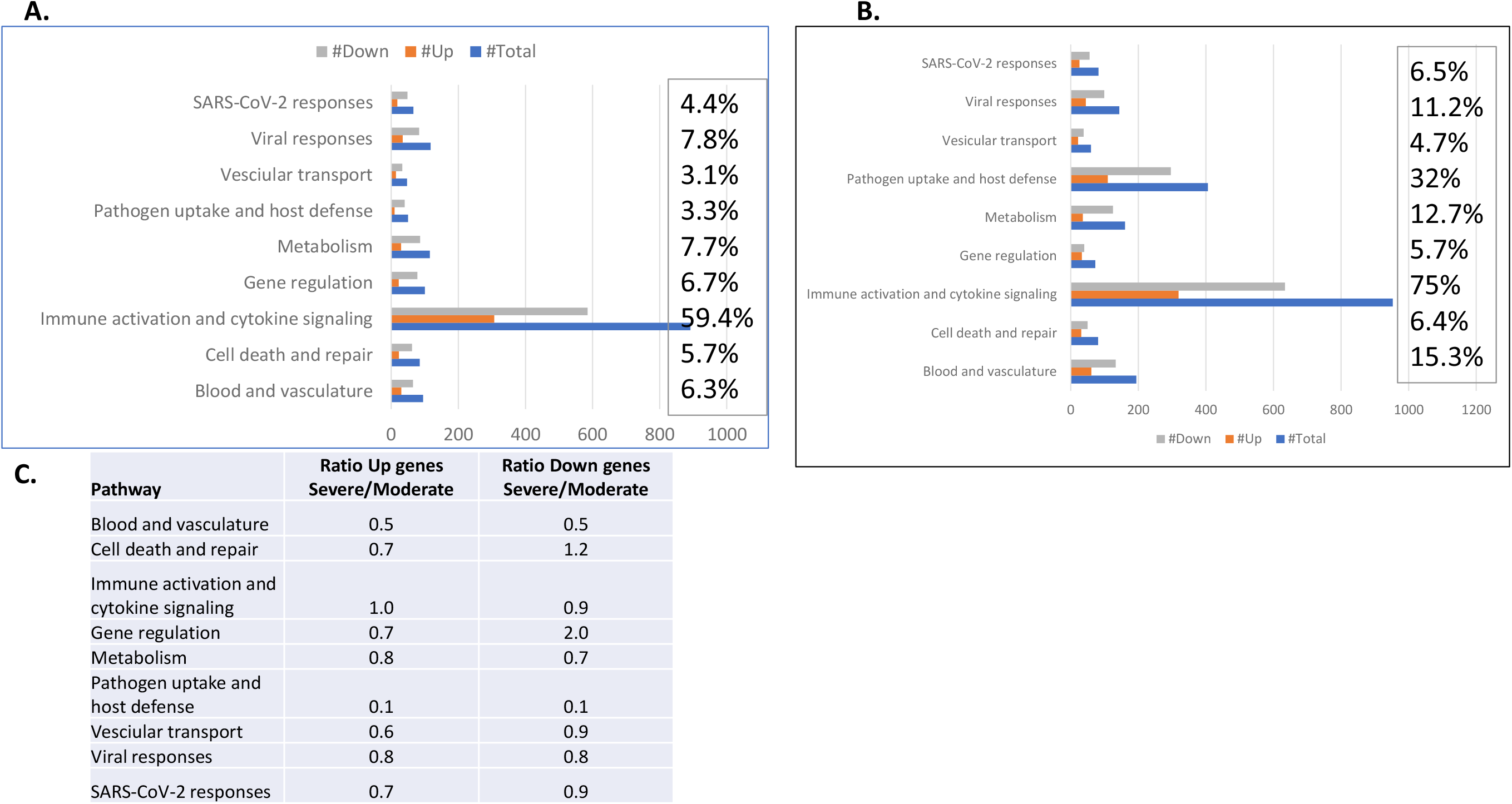
Cellular pathways differentially regulated in COVID-19 cases. A. EC and moderate Covid cases, B. EC and Severe, Up list is raised in EC and Down list is raised in Covid. C, Gene ratios found between Up and Down gene lists for ‘B’ and ‘C’ in particular pathway sub-groups based on DEGs in Severe/Moderate Covid.

Immune activation and cytokine signaling pathway genes with 307 Up and 585 Down genes included, Interleukin 1, −2,−6, −4,−5, 10, 12,−13, TNFα and NFKappa-b pathways, Interferon-7alpha/beta signaling, T and B cell receptor activation and intracellular signaling such as, mitogen activated protein kinase (MAPK), MyD88-independent toll like receptor (TLR) cascade and cell surface receptor interactions such as TLRs and integrin genes (Supplementary Table 1). Pathways related to complement activation, receptor binding, triggering of host defenses through antimicrobial peptides and defensins together with MMP were differentially activated between controls and COVID-19 cases.

There were 29 Up and 86 Down regulated linked to glucose, insulin pathways and the hypoxia stress response. We found 30 Up and 65 Down genes belonging to blood clotting, platelet responses, endothelin pathway, fibrin clotting and vascular interactions. Cell death and repair genes (23 Up and 62 Down) belonged to, the apoptotic response, TP53 and DNA damage response pathways. DEGs belonging to vesicular transport such as, clathrin-mediated endocytosis, Golgi-transport and actin cytoskeletal regulation were differentially regulated in COVID-19.

Of the known SARS-CoV-2 induced pathways, we found 18 Up and 48 Down genes between EC and moderate COVID-19. SARS-CoV-2 infection induced-MAPK, interferon signaling, apoptosis, ER stress, innate immune activation, NRLP3 inflammasome responses, ACE2 Receptor pathway, autophagy and hijack of ubiquitination related genes were upregulated in COVID-19. Additional responses pathways previously shown to be induced by Ebola virus, HIV and Influenza were identified including 16 genes upregulated and 35 downregulated in COVID-19.

DEGs in severe COVID-19 as compared with ECs revealed the largest sub-group to belonged to immune activation and cytokine signaling pathways (75%), followed by pathogen uptake and host defenses (32%), blood and vasculature (15%), glucose metabolism (13%), viral induced pathways (11.3), SARS-CoV-2 responses (6.5%), cell death and repair (6.4%), gene regulation (5.7%) and vesicular transport (4.7%), Fig. 2B, Supplementary Table 2.

**Table 2.**
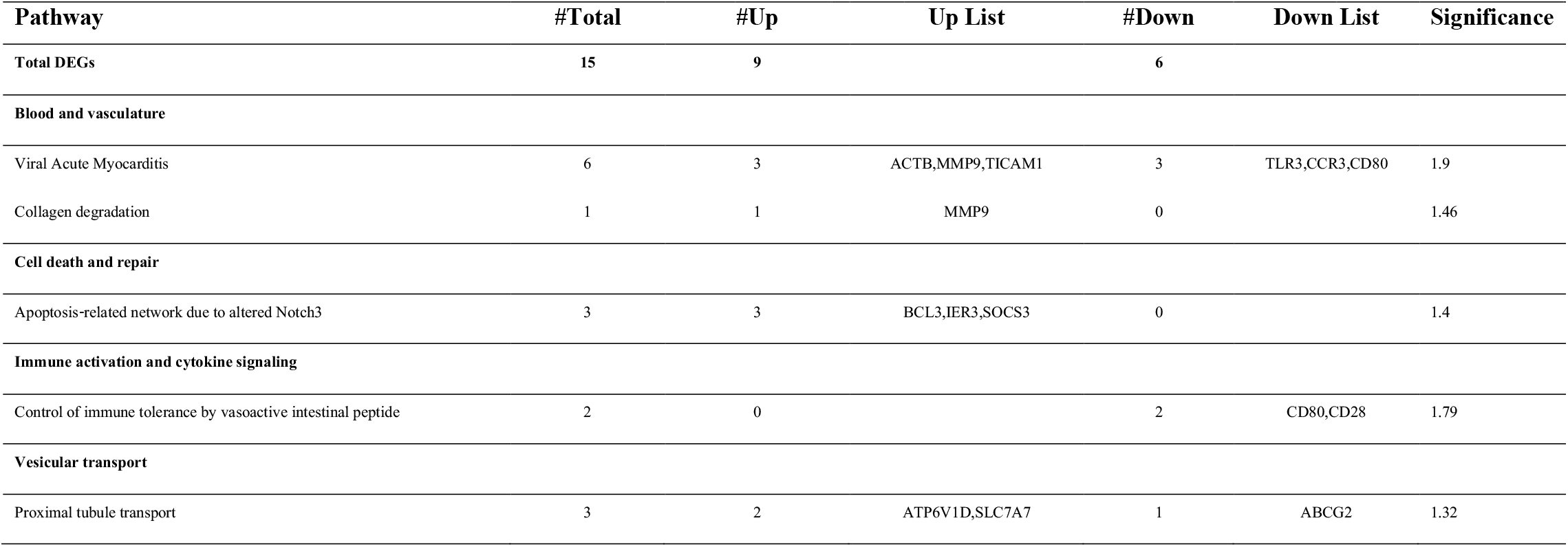
DEGs in COVID-19 cases with critical as compared with moderate disease.

| **Pathway** | **#Total** | **#Up** | **Up List** | **#Down** | **Down List** | **Significance** |
| --- | --- | --- | --- | --- | --- | --- |
| **Total DEGs** | **15** | **9** |  | **6** |  |  |
| **Blood and vasculature** |  |  |  |  |  |  |
| Viral Acute Myocarditis | 6 | 3 | ACTB,MMP9,TICAM1 | 3 | TLR3,CCR3,CD80 | 1.9 |
| Collagen degradation | 1 | 1 | MMP9 | 0 |  | 1.46 |
| **Cell death and repair** |  |  |  |  |  |  |
| Apoptosis-related network due to altered Notch3 | 3 | 3 | BCL3,IER3,SOCS3 | 0 |  | 1.4 |
| **Immune activation and cytokine signaling** |  |  |  |  |  |  |
| Control of immune tolerance by vasoactive intestinal peptide | 2 | 0 |  | 2 | CD80,CD28 | 1.79 |
| **Vesicular transport** |  |  |  |  |  |  |
| Proximal tubule transport | 3 | 2 | ATP6V1D,SLC7A7 | 1 | ABCG2 | 1.32 |

Upon comparison of DEGs between moderate and severe COVID-19 cases, we found those related to blood and vasculature with a 50% reduction in the number of modulated in severe as compared with moderate cases. In pathogen uptake and host defense DEGs, there was 10% of the change in DEGs in severe as compared with moderate cases (Fig. 2C).

Comparison of the RNA transcriptomes severe and moderate COVID-19 cases revealed that 268 DEGs, with 110 upregulated and 158 downregulated in severe cases, Fig. 3A-C. In particular, genes related to viral acute mycocarditis, ACTB, MMP9, TICAM1 were upregulated whilst TLR3, CCR3 and CD80 were downregulated (Table 2). Apoptosis related network genes BLC3, IER3 and SOCS3 were upregulated in severe cases. Immune tolerance genes CD80 and CD28 were downregulated in severe cases.

**Fig 3.**
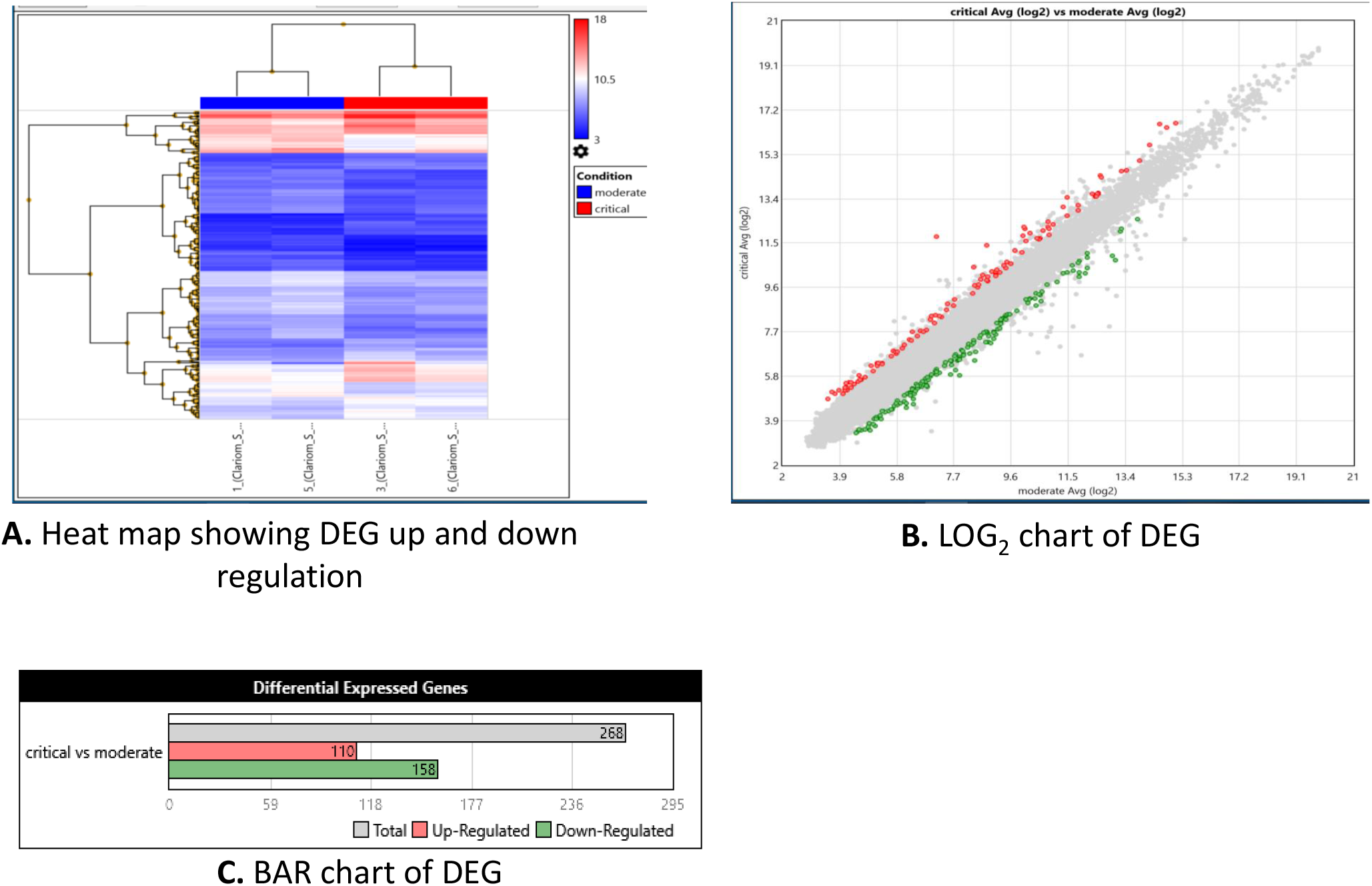
Differential regulation between severe and moderate COVID-19. **A**. Heat map showing hierarchical clustering of DEGs between severe and moderate cases B. Volcano plot showing Log2 difference between DEGs in severe and moderate disease. **C**, Histogram showing DEGs either Up or Down between groups.

### DEGs related to unfavourable disease progression in critical COVID-19

We compared transcriptional signatures at 3 and 6 days after admission in case P3, who subsequently expired. We observed 7693 DEGs between the two time points, whereby 5151 were upregulated and 2542 were downregulated compared between the time points. The DEGs primarily involved immune regulation and cytokine signaling, blood and vasculature, SARS-CoV-2 and viral induced responses, cell death and repair, pathogen uptake and host defense pathways (Fig. 4). Blood and vasculature related pathway genes were downregulated through disease progression included genes related to cell surface interactions at the vascular surface such as, VCAM1, platelet homeostasis and cardiac conduction related genes such as, CALM1, blood and fibrin clotting pathway genes (SERPINs) and endothelin pathway genes.

**Fig 4.**
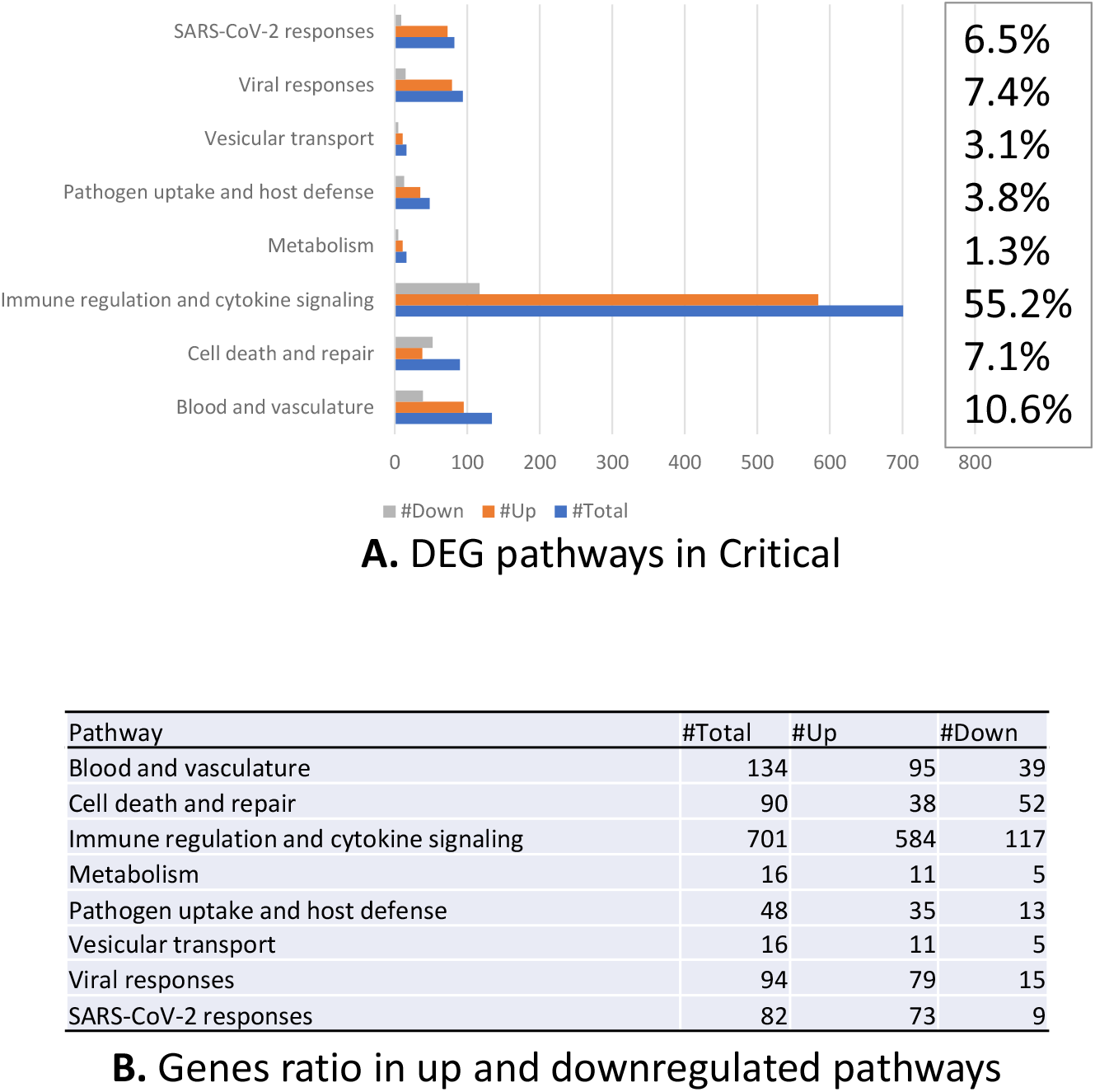
DEGs in a critical COVID-19 case with disease progression. **A**. Histogram showing DEGs a case with severe/critical disease at 3 and 6 days of admission in hospital B. Ratios of genes Up or Down regulated between 3 and 6 days.

Downregulated genes included cellular signaling such as, MAPK, interleukin IL-2, −10, IFNα/β, TLR3, Myd88 independent TLR cascade and genes involved in selective expression of chemokine receptors involved in T cell polarization (Supplementary Table 3). Further, SARS-CoV-2 induced Type 1 interferon response, MAPK pathway and human CoV related apoptosis pathway, autophagy and ubiquitination response genes were all reduced with time in the critical COVID-19 patient.

## Discussion

The COVID-19 disease spectrum of ranges from asymptomatic, mild upper respiratory tract symptoms to severe and critical disease associated with a cytokine release syndrome that leads to hypoxia in the lung and acute respiratory failure (9). While SARS-CoV-2 infection is mostly thought to be a threat for elderly individuals and those with multiple co-morbid conditions (10), in children COVID-19 has been associated with an acute inflammatory syndrome similar to Kawaski’s disease or those related to reactivation of viral infections (11). Further, COVID-19 is associated with thrombosis and increased risk of strokes (1). Providing effective treatment and management of COVID-19 requires understanding of disease progression and risk that can identify potential therapeutic interventions. Here we investigated host transcriptome responses in patients with moderate and severe COVID-19 to understand mechanisms which are dysregulated during SARS-CoV-2 infection.

The DEGs genes most affected in COVID-19 cases in our study were found to be related to immune activation and cytokine signaling pathways. Key inflammatory genes including, Toll like receptor (TLR), chemokine signaling, JAK/STAT, IL-1,2, 3, 5, GM-CSF and MAPK signaling pathways were increased (Sup Tables 1 and 2). We found, SARS-CoV-2 specific IFN I along with IL10 and TGFβ were found to be up-regulated in COVID-19 cases as compared with the controls. The expression of pro-inflammatory cytokines and chemokines are necessary for initial clearance of viral infection, however, the aggravated expression of these have been linked with excessive tissue damage leading to the development of acute respiratory distress syndrome (ARDS) (12). Previous reports have shown that in SARS effected patients, poor prognosis is associated with severe respiratory failure (13). ARDS is shown to be associated with the hyper-immune activation in patients with SARS leading to severe tissue damage and hypoxia (14).

These results corroborate with previous reports that viral infections modulate host machinery to dysregulate host immune responses and support replication and spread (15, 16). SARS-CoV2 potentiates its effect mostly through triggering of a ‘cytokine storm’ with devastating consequences on host inflammation (17). SARS-CoV2 infection can cause severe pulmonary disease with complications including extensive tissue damage, blood clots formation leading to stroke in adults (18).

Patients with severe COVID-19 had compromised lung function which was evident by neutrophil accumulation, increased lung edema and the need to be supported by mechanical ventilation. Genes related to endothelial and vascular interactions were differentially regulated in the cohort of severely ill patients as compared to the healthy population. The progressive lung damage witnessed in the severe cases of COVID-19 also results from hyper-active inflammatory immune response. The gene expression signature in response to SARS-CoV-2 reported has shown that COVID 19 disease is marked by uncontrolled production of inflammatory markers being referred to as cytokine storm (6, 19). The viral response genes that were effected in the cohort of severely ill patients were decreased Angiotensin converting enzyme 2 (ACE2) pathway gene TMPRSS2 expression with increased NOD-like receptor protein 3 (NLRP3) inflammasome, together with oxidative stress genes, cell death and repair genes including apoptotic response and TP53 pathways and DNA damage response pathways (STable 2). SARS-CoV-2 enters into the host cell through ACE 2 receptors (20) and triggers the activation of NLRP3 inflammasome which further releases inflammatory cytokines (21). Up-regulated inflammatory cytokines such as IL1β and TNFα has also shown to play a role in ACE2 shedding (22, 23). Loss of pulmonary ACE2 function has been suggested to be associated with acute lung injury (24). We also found DEGs related to oxidative stress to be raised in our COVID-19 cases. Accumulated neutrophils release histo-toxic mediators comprising of reactive oxygen species (ROS) causing cellular damage which could be responsible for this. Inflammation caused leads to increased gaps in the endothelium resulting in endothelial dysfunction associated with apoptosis (25, 26).

DEGs in severe COVID-19 cases were related to blood clotting, platelet homeostasis, cell surface interactions with blood vascular surfaces and the endothelin pathway. Breakdown products of fibrin, D-dimers are a biomarker of inflammation and progressive disease in COVID-19 and these were increased in all our COVID-19 study subjects. Endothelial cells are sensitive to oxidative stress and can be damaged due to an imbalance resulting from inflammatory cytokine related pathways (26). Histological analysis of tissues from COVID-19 cases has shown endothelial lining to disrupted in lung tissue (26). Therefore, worsening COVID-19 may be due to SARS-CoV-2 affecting endothelial cell driven responses, coagulopathy and blood clotting related disturbances. We found the cardiac conduction related genes such as, CALM1 to be upregulated in COVID-19 cases. CALM1 encodes for calmodulin which plays an essential role in cardiac contraction. We found SERPINB2 to be upregulated in blood and fibrin clotting pathway and endothelin pathway genes (Sup Table2). SERPINB2 or plasminogen activator-2 is coagulation factor that inhibits tissue plasminogen activator. Upregulation of SERPINB2 increases the risk of thrombosis. Therefore, upregulation of SERPINB2 may be a mechanism which induces in the increased risk of thrombosis in COVID-19 patients.

Despite the upregulated IFN genes, the downstream TCR and co-stimulatory signaling pathway were down-regulated in patients with COVID-19 as compared with healthy controls. T cells effector responses are down-regulated in COVID-19 cases whilst, a slightly increase in T cells has been recorded post treatment (27). The suppression of immune activation and cytokine genes in SARS-CoV2 infected patients may lead to defective clearance of viral infection. SARS-CoV-2 has been shown to induce reduced IFN type I responses, increased pro-inflammatory cytokines and chemokines profile with down-regulated IL-10 responses (6, 19). Expression of IFNs has been shown to be essential in viral infection as in addition to upregulating anti-viral immune responses in APCs, IFN also mediates adaptive immune responses by activating T cells. The balance in which IFN responses are required is controlled by immune modulatory cytokines such as IL10 and TGFβ to prevent the tissue damage (28).

On comparison of severe and moderate COVID-19 cases we found that genes related to viral acute myocarditis (MMP9, TICAM1) were upregulated whilst TLR, CCR3 and CD80 were downregulated. MMP9 is associated with inflammation in the host whilst TLR mediated pathways are induced in SARS-COV-2 host immune activation (29). CD80 binds CD28 and plays a role in lymphocyte activation and signaling, its downregulation likely suggests reduction of adaptive immunity in the host. Study of SARS-CoV2 infected host transcriptomes has revealed that the virus activates a range of immune activation and cytokine signaling pathways, those which are related to apoptosis and cell cycle regulation, and modifies host defense responses trigger by pathogen uptake and internalization (19). Recent reports have shown that SARS-CoV2 exhibits some similar virogenomic signatures of pathogenic viruses such as Ebola, SARS, H1N1 and MERS (Middle East respiratory syndrome virus) including the plasminogen activator (SERPINB1) but also distinct immune inflammatory signatures (30).

## Conclusions

Our study indicates indicates a down regulation of immune regulation and cytokine signaling with disease progression and dysregulation of genes related to blood and vasculature followed by pathogen uptake and host defense pathways. Genes involved in fibrin clotting, platelet homeostasis and endothelin pathways were all affected in COVID-19. Treatment of patients with interventions for these patients such as anti-coagulation therapy in addition to anti-inflammatory agents may improve treatment strategies for COVID-19.

## Data Availability

Supplementary files for this manuscript will be available after final publication of the manuscript.

## Acknowledgements

This study received support through a University Research Council grant, The Aga Khan University and the Higher Education Commission, Pakistan. Thanks to the Clinical Laboratory team, The Aga Khan University Hospital, Dr. Zeeshan Ansar and Nazneen Islam for laboratory support and to Maliha Yameen for technical assistance.

## References

1. Guzik TJ, Mohiddin SA, Dimarco A, Patel V, Savvatis K, Marelli-Berg FM, et al. COVID-19 and the cardiovascular system: implications for risk assessment, diagnosis, and treatment options. Cardiovasc Res. 2020.

2. Docherty AB, Harrison EM, Green CA, Hardwick HE, Pius R, Norman L, et al. Features of 20 133 UK patients in hospital with covid-19 using the ISARIC WHO Clinical Characterisation Protocol: prospective observational cohort study. BMJ. 2020;369:m1985.

3. Zhou Y, Hou Y, Shen J, Huang Y, Martin W, Cheng F. Network-based drug repurposing for novel coronavirus 2019-nCoV/SARS-CoV-2. Cell Discov. 2020;6:14.

4. Walls AC, Park YJ, Tortorici MA, Wall A, McGuire AT, Veesler D. Structure, Function, and Antigenicity of the SARS-CoV-2 Spike Glycoprotein. Cell. 2020;181(2):281–92 e6.

5. Park WB, Kwon NJ, Choi SJ, Kang CK, Choe PG, Kim JY, et al. Virus Isolation from the First Patient with SARS-CoV-2 in Korea. J Korean Med Sci. 2020;35(7):e84.

6. Fung SY, Yuen KS, Ye ZW, Chan CP, Jin DY. A tug-of-war between severe acute respiratory syndrome coronavirus 2 and host antiviral defence: lessons from other pathogenic viruses. Emerg Microbes Infect. 2020;9(1):558–70.

7. Qin C, Zhou L, Hu Z, Zhang S, Yang S, Tao Y, et al. Dysregulation of immune response in patients with COVID-19 in Wuhan, China. Clin Infect Dis. 2020.

8. Tay MZ, Poh, C.M., Rénia, L. et al. The trinity of COVID-19: immunity, inflammation and intervention. Nat Rev Immunol. 2020.

9. Guan WJ, Ni ZY, Hu Y, Liang WH, Ou CQ, He JX, et al. Clinical Characteristics of Coronavirus Disease 2019 in China. N Engl J Med. 2020;382(18):1708–20.

10. Meisner BA, Boscart V, Gaudreau P, Stolee P, Ebert P, Heyer M, et al. Interdisciplinary and Collaborative Approaches Needed to Determine Impact of COVID-19 on Older Adults and Aging: CAG/ACG and CJA/RCV Joint Statement. Can J Aging. 2020:1–31.

11. Viner RM, Whittaker E. Kawasaki-like disease: emerging complication during the COVID-19 pandemic. Lancet. 2020.

12. Pugin J, Ricou B, Steinberg KP, Suter PM, Martin TR. Proinflammatory activity in bronchoalveolar lavage fluids from patients with ARDS, a prominent role for interleukin-1. Am J Respir Crit Care Med. 1996;153(6 Pt 1):1850–6.

13. Lew TW, Kwek TK, Tai D, Earnest A, Loo S, Singh K, et al. Acute respiratory distress syndrome in critically ill patients with severe acute respiratory syndrome. JAMA. 2003;290(3):374–80.

14. Wang D, Hu B, Hu C, Zhu F, Liu X, Zhang J, et al. Clinical Characteristics of 138 Hospitalized Patients With 2019 Novel Coronavirus-Infected Pneumonia in Wuhan, China. JAMA. 2020.

15. Xu LH, Huang M, Fang SG, Liu DX. Coronavirus infection induces DNA replication stress partly through interaction of its nonstructural protein 13 with the p125 subunit of DNA polymerase delta. J Biol Chem. 2011;286(45):39546–59.

16. Channappanavar R, Fehr AR, Vijay R, Mack M, Zhao J, Meyerholz DK, et al. Dysregulated Type I Interferon and Inflammatory Monocyte-Macrophage Responses Cause Lethal Pneumonia in SARS-CoV-Infected Mice. Cell Host Microbe. 2016;19(2):181–93.

17. Zeng F, Huang Y, Guo Y, Yin M, Chen X, Xiao L, et al. Association of inflammatory markers with the severity of COVID-19: a meta-analysis. Int J Infect Dis. 2020.

18. Ullah W, Saeed R, Sarwar U, Patel R, Fischman DL. COVID-19 complicated by Acute Pulmonary Embolism and Right-Sided Heart Failure. JACC Case Rep. 2020.

19. Xiong Y, Liu Y, Cao L, Wang D, Guo M, Jiang A, et al. Transcriptomic characteristics of bronchoalveolar lavage fluid and peripheral blood mononuclear cells in COVID-19 patients. Emerg Microbes Infect. 2020;9(1):761–70.

20. Fu Y, Cheng Y, Wu Y. Understanding SARS-CoV-2-Mediated Inflammatory Responses: From Mechanisms to Potential Therapeutic Tools. Virol Sin. 2020.

21. Chen IY, Moriyama M, Chang MF, Ichinohe T. Severe Acute Respiratory Syndrome Coronavirus Viroporin 3a Activates the NLRP3 Inflammasome. Front Microbiol. 2019;10:50.

22. Jia HP, Look DC, Tan P, Shi L, Hickey M, Gakhar L, et al. Ectodomain shedding of angiotensin converting enzyme 2 in human airway epithelia. Am J Physiol Lung Cell Mol Physiol. 2009;297(1):L84–96.

23. Lambert DW, Yarski M, Warner FJ, Thornhill P, Parkin ET, Smith AI, et al. Tumor necrosis factor-alpha convertase (ADAM17) mediates regulated ectodomain shedding of the severe-acute respiratory syndrome-coronavirus (SARS-CoV) receptor, angiotensin-converting enzyme-2 (ACE2). J Biol Chem. 2005;280(34):30113–9.

24. Imai Y, Kuba K, Rao S, Huan Y, Guo F, Guan B, et al. Angiotensin-converting enzyme 2 protects from severe acute lung failure. Nature. 2005;436(7047):112–6.

25. Pober JS, Sessa WC. Evolving functions of endothelial cells in inflammation. Nat Rev Immunol. 2007;7(10):803–15.

26. Varga Z, Flammer AJ, Steiger P, Haberecker M, Andermatt R, Zinkernagel AS, et al. Endothelial cell infection and endotheliitis in COVID-19. Lancet. 2020;395(10234):1417–8.

27. Ouyang Y, Yin J, Wang W, Shi H, Shi Y, Xu B, et al. Down-regulated gene expression spectrum and immune responses changed during the disease progression in COVID-19 patients. Clin Infect Dis. 2020.

28. Moss RB, Moll T, El-Kalay M, Kohne C, Soo Hoo W, Encinas J, et al. Th1/Th2 cells in inflammatory disease states: therapeutic implications. Expert Opin Biol Ther. 2004;4(12):1887–96.

29. Li G, Fan Y, Lai Y, Han T, Li Z, Zhou P, et al. Coronavirus infections and immune responses. J Med Virol. 2020;92(4):424–32.

30. Alsamman AMZ, H;. The transcriptomic profiling of COVID-19 compared to SARS, MERS, Ebola, and H1N1. bioRxiV. 2020(https://doi.org/10.1101/2020.05.06.080960)M. Alsamman, View ORCID ProfileHatem Zayed.

